# Modelling long COVID using Bayesian networks

**DOI:** 10.1101/2024.03.04.24303715

**Authors:** Gladymar Pérez Chacón, Steven Mascaro, Marie J. Estcourt, Chan Phetsouphanh, Ann E. Nicholson, Tom Snelling, Yue Wu

**Author notes:** Corresponding author *Email address:* (Yue Wu). The Kirby Institute, University of New South Wales, Kensington, New South Wales, Australia (Gladymar Pérez Chacón). Also with Bayesian Intelligence, Upwey, Melbourne, Australia (Steven Mascaro). Also with Wesfarmers Centre of Vaccines and Infectious Diseases, Telethon Kids Institute, Nedlands, Western Australia, Australia (Tom Snelling).

## Abstract

Motivated by the ambiguity of operational case definitions for long COVID and the impact of the lack of a common causal language on long COVID research, in early 2023 we began developing a research framework on this post-acute infection syndrome. We used directed acyclic graphs (DAGs) and Bayesian networks (BNs) to depict the hypothesised mechanisms of long COVID in an agnostic fashion. The DAGs were informed by the evolving literature and subsequently refined following elicitation workshops with domain experts. The workshops were structured online sessions guided by an experienced facilitator. The causal DAGs aim to summarise the hypothesised pathobiological pathways from mild or severe COVID-19 disease to the development of pulmonary symptoms and fatigue over four different time points. The DAG was converted into a BN using qualitative parametrisation. These causal models aim to assist the identification of disease endotypes, as well as the design of randomised controlled trials and observational studies. The framework can also be extended to a range of other post-acute infection syndromes.

## 1. Introduction

Between 2020 and 2021, the estimated global prevalence of self-reported fatigue, respiratory symptoms, and cognitive decline spanning at least 3 months from the onset of SARS-CoV-2 infection was 6.2% (95% uncertainty interval [UI], 2.4%-13.3%).[1] The low precision of the UI reflects heterogenous and limited data which hamper our understanding of the series of complex conditions that can arise after the acute phase of COVID-19; these conditions are generally referred to in the literature as post-acute sequelae of COVID, or ‘long COVID’.

SARS-CoV-2 reservoirs,[2] tissue damage and dysfunction triggered by inflammation,[3] immune dysregulation,[4] mitochondrial dysfunction,[5], and reactivation of Epstein-Barr virus (EBV) or other latent viruses [6] have been hypothesised as pathobiological drivers of long COVID. However, in order to answer (quantitative) causal questions about long COVID, we need to understand the causal pathways that link long COVID symptoms to their proposed underpinning mechanisms.[3] This effort has been stymied by the lack of a common language and the diversity of long COVID definitions.

To overcome these obstacles, here we present a research framework based on a causal modelling approach that harmonises both operational case definitions and proposed pathological mechanisms of long COVID using causal directed acyclic graphs (DAGs) and Bayesian networks (BNs). A DAG is a graphical model that depicts assumptions about the relationship between nodes (variables) using arcs (arrows) which go from one node to the other, with no directed cycles. A BN is a DAG in which the dependencies between the nodes are represented by a series of conditional probability distributions (Figure 1).[7] Therefore, a BN allows causal reasoning in an uncertain domain, based on a graphical structure (a DAG) coupled with associated conditional probability estimates.[7]

**Figure 1:**
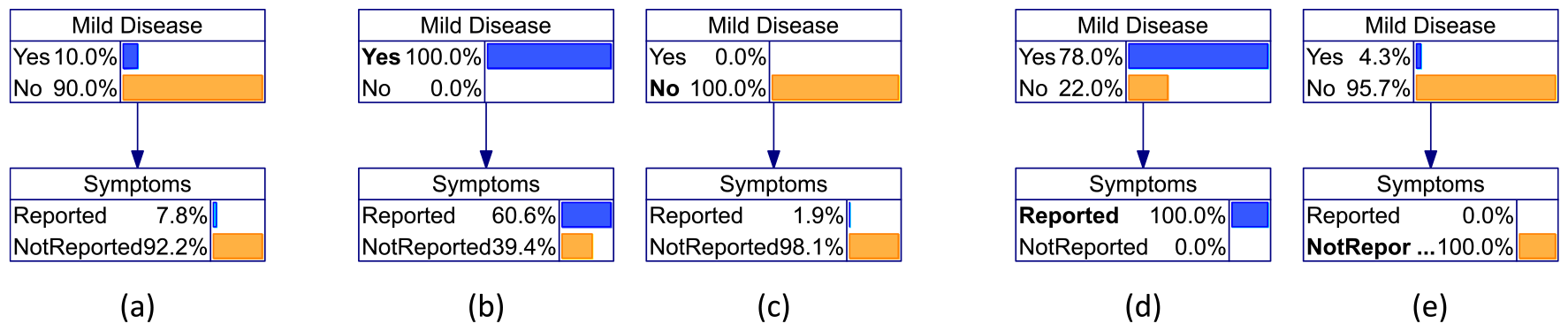
Simple two-node example of a Bayesian network describing the probabilistic relationship between disease and symptoms with variation in the availability of evidence. Here we show probabilities for all nodes when there is: (a) no evidence; evidence of (b) mild disease or (c) no mild disease; evidence of (d) reported symptoms or (e) symptoms not reported.

The nodes in a BN depict a set of random variables from the domain. The arcs (or arrows) represent direct (unmediated) causal dependencies when used to encode causal knowledge.[7] Assuming discrete variables, the direct causal dependencies are quantified by conditional probability tables associated with each node. This encoding allows one to identify potential issues with statistical inference ahead of an analysis (such as contextual and definitional differences, as well as the impact from confounding, selection bias, and measurement error) and opportunities for resolving them.[8] The predictions of a BN can be updated along the patient’s clinical course, utilising as little or as much information as may become available over time,[7] making them a good option for decision support in settings where information may be evolving and/or sparse.

BNs can be used to model systems that change over time (e.g., dynamic systems) using sequential data. These “dynamic” BNs are computationally complex state-space models which aim to learn and accurately represent the problem domain of interest.[9] This can help facilitate discussion among scientific researchers, medical experts, and people with lived experience. Likewise, they may ultimately assist us in better understanding who is more likely to develop long COVID and their pathways to recovery. These features allow studies conducted under different conditions to be compared and integrated.

## 2. Results

### 2.1. A general framework on post-COVID phenomena

The general framework (Figure 2) models the natural history of SARS-CoV-2 infection across a prolonged period of time (*t*) from *t*_1_ (the start of acute infection) to *t*_4_ (e.g., 2 years post-infection). We broadly divide COVID-19 into mild (e.g., ambulatory, **M1**) and severe (e.g., hospitalised, **V1**) cases during the acute phase of infection, where a mild case can progress to become severe. Both mild (**M1**) and severe (**S1**) COVID-19 can cause reversible organ dysfunction (**R1**) or persistent organ damage and dysfunction (**P1**). In the first instance, reversible organ injury is followed by recovery. When organ injury is severe, homeostatic self-repair may fail and the organ injury will persist into future time periods. Any reversible (**R1**) or persistent (**P1**) organ dysfunction may manifest as patient-reported symptoms (**S1**). Different pathobiological mechanisms causing organ dysfunction can give rise to similar symptoms. It is important to note that the severity of specific organ damage and dysfunction may fluctuate over time. As a result, the presence of symptoms caused by the same underlying mechanism can also fluctuate (e.g., recurrent pattern, **S3**), which may pose challenges for data collection.

**Figure 2:**
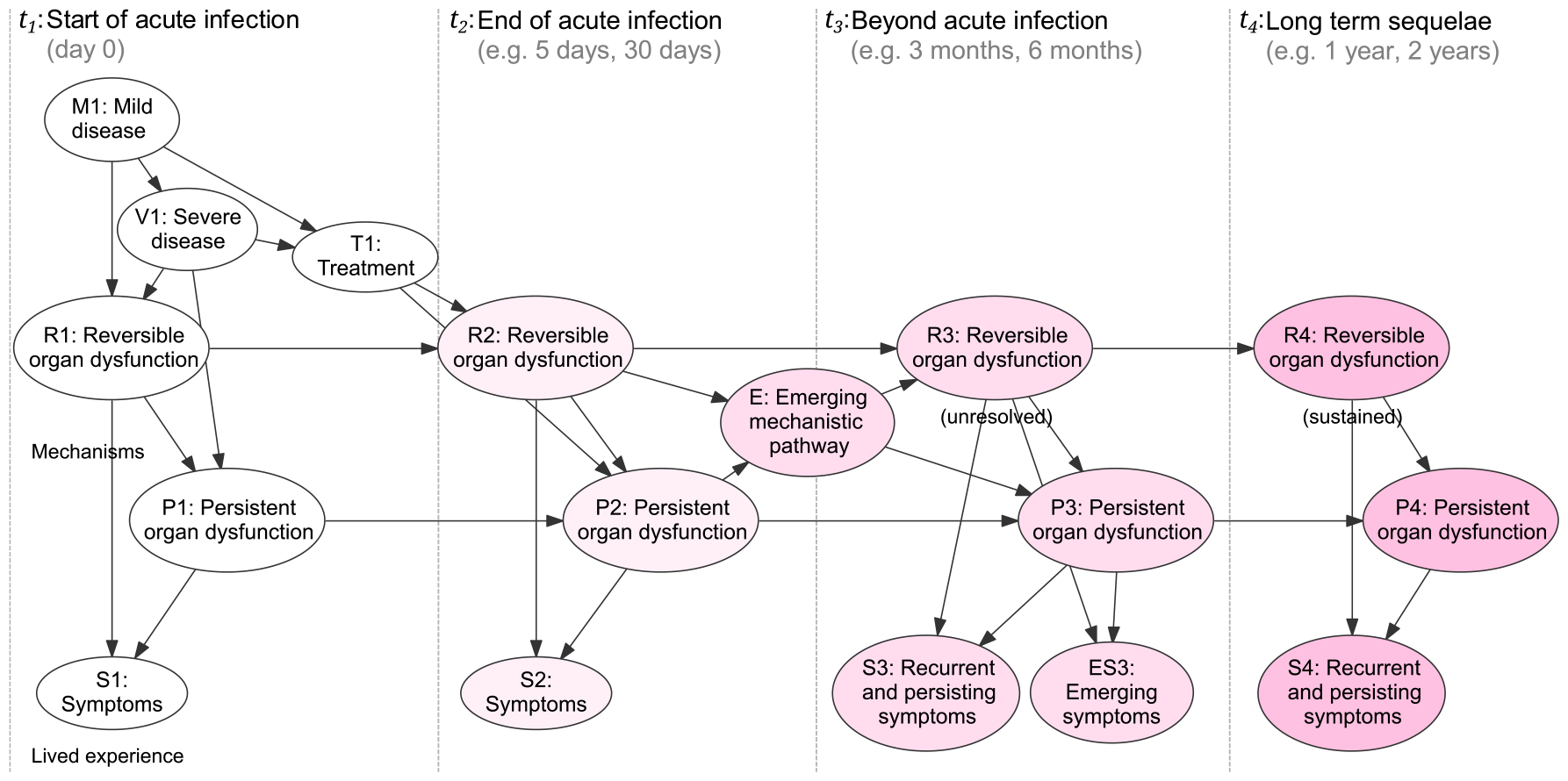
Qualitative dynamic causal Bayesian network that represents the natural history of long COVID (which is referred to in the text as the general framework). Four different time periods of interest were chosen, defined by the start of acute infection (*t*_1_), which can be confirmed by, e.g., PCR, suspected on the basis of less direct evidence, or otherwise remain latent; the end of the acute infection (*t*_2_); beyond acute infection (*t*_3_, e.g., 3 months post *t*_1_), and long-term sequelae (*t*_4_, e.g., 1-to-2 years post *t*_1_). This dynamic model encompasses six variables at *t*_1_, identified with a capital letter (**M1**, mild disease; **V1**, severe disease; **T1**, treatment; **R1**, reversible organ dysfunction; **P1**, persistent organ dysfunction; **S1**, symptoms); three of them (**R, I**, and **S**) can further change their status throughout the follow-up period (*t*_2_ to *t*_4_). An emerging mechanistic pathway (**E**) can arise at any time point. In this general framework, **E** is the child of **R2** and **P2**, and can directly influence the development of unresolved and sustained dysfunctions at *t*_3_. Emerging mechanisms may also lead to emerging symptoms (**ES3**). For simplicity, no background factors are included in the models.

Guided by any clinical indicators of the disease status (mild [**M1**] or severe [**V1**] COVID-19), the acute infection may be treated (**T1**), leading to an updated functional status for relevant organs at the end of acute infection (*t*_2_). Under the circumstance where organ dysfunction continues beyond the acute phase (passing from *t*_2_ into *t*_3_ and *t*_4_), symptoms could persist or recur (**S3** and **S4**). As multiple organs interact as a (dynamic) system, new mechanistic pathways (**E**) may emerge and give rise to organ dysfunction that was not previously present, which may subsequently give rise to new symptoms (**ES3**). New symptoms may also appear due to a worsening of pre-existing organ dysfunction (**R3** or **P3**).

### 2.2. Illustrative scenarios

The general framework was qualitatively parameterised in order to assess its ability to represent the expected patterns of disease progression — that is, with the sole concern of capturing the intended qualitative behaviour, including the direction and approximate size of probability changes in response to new evidence (Figure 3).[10] Supported by this qualitative parameterisation, we present four scenarios to illustrate and test the use of the general framework on post-COVID phenomena. The test here is solely that the *causal structure* is able to represent broad classes of expected model behaviour; the scenarios require - but are not a test of - the qualitative parameterisation. The states of the input variables for each scenario are characterised as *yes* or *reported* and are therefore considered to be ‘observed evidence’ which may be an imperfect measure of the concept in question (for example, someone may fail to report presence of cough, or it may be incorrectly reported as present when it is not). The four scenarios are intended to be broadly illustrative for any era or strain of SARS-CoV-2, but otherwise assume natural infection in the absence of immunity from vaccination or prior infection. In the following subsections, probabilities have all been rounded to the nearest whole probability.

**Figure 3:**
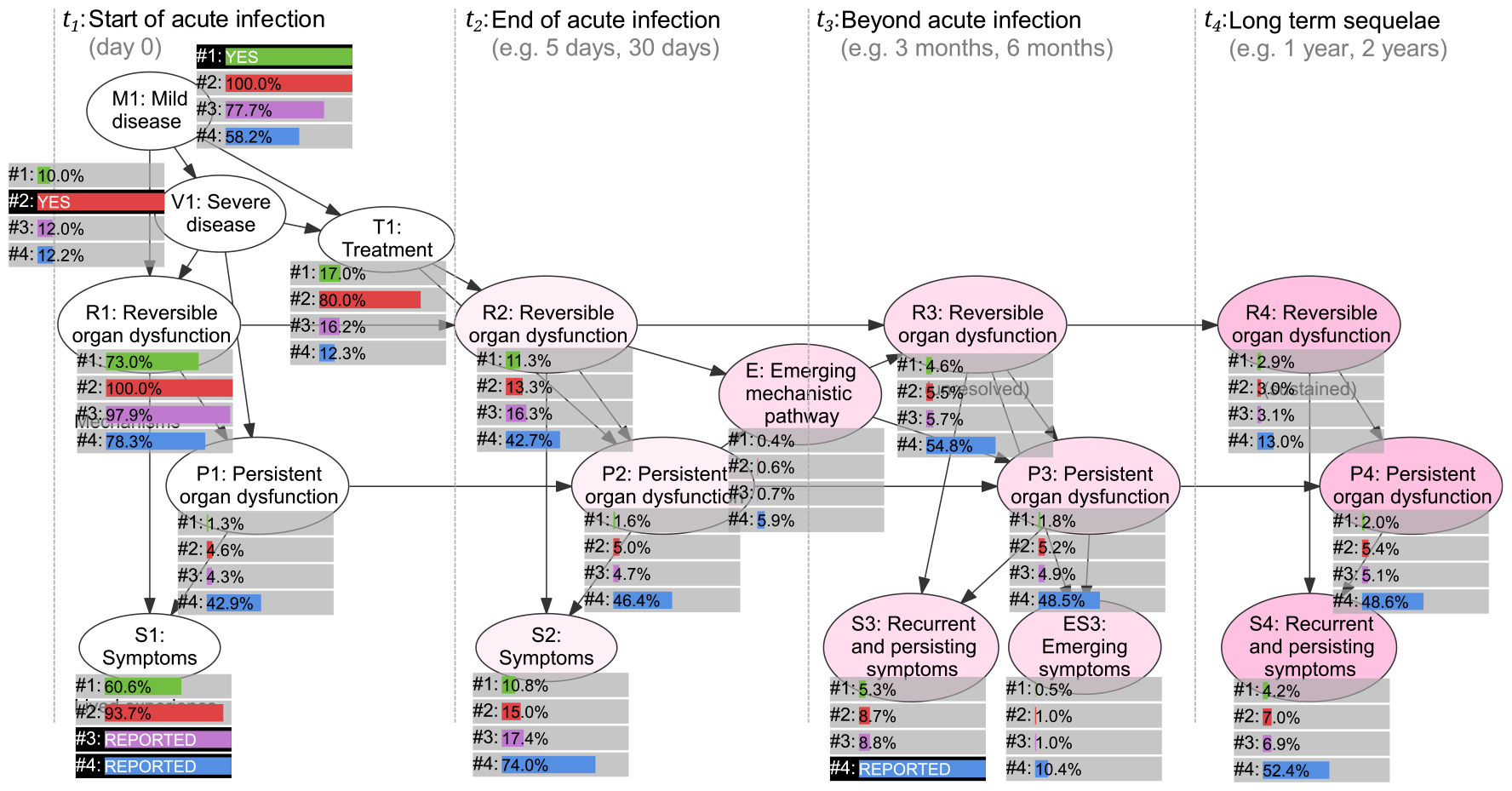
Four scenarios (labelled #1, #2, #3, or #4 on each node) with the evidence and probabilities for each node in the scenario. When a node has evidence entered for it in a given scenario, the evidence is shown with a black background; otherwise, the probability for that node is shown, along with a bar, coloured differently for each scenario (green, red, purple and blue, respectively). In scenario 1, there is one piece of evidence, that **M1: Mild disease** is *yes*. In scenario 2, the evidence instead is that **V1: Severe disease** is *yes*. In scenario 3, the only evidence is that **V1: Symptoms** are *reported* at *t*_1_. In scenario 4, there are two pieces of evidence, that symptoms are *reported* at *t*_1_ (**S1: Symptoms**) and *t*_3_ (**S3: Symptoms**). Across the four scenarios, the probability of **P1: Persistent organ dysfunction** being true, for example, is 1.3% (scenario 1), 4.6% (scenario 2), 4.3% (scenario 3) and 42.9% (scenario 4).

#### 2.2.1. Scenario 1: Mild disease observed at t_1_

Consider an adult with observed mild COVID-19 (**M1**) at *t*_1_. The model depicts a 10% probability of progressing to severe disease (**V1**), which gives rise to a 73% probability of developing reversible organ dysfunction (**R1**), and a very small probability (1%) of persistent organ dysfunction (**P1**) at *t*_1_. The mechanisms are not directly observable; what may be observed is the 61% probability of reporting symptoms (**S1**, e.g., **cough** or **fatigue**). The probability of sustained reversible organ dysfunction drops to 11% at *t*_2_, 5% at *t*_3_ and 3% at *t*_4_, as the underlying tissue damage and dysfunction recovers. Meanwhile, persistent organ dysfunction (**P1**), by dint of its persistence, will rarely decrease in probability — here increasing from 1% to 2% at *t*_4_. The probability of reported symptoms would also drop from the initial 61% (driven by reversible dysfunction [**R1**]) to 4% over the modelled period (**S4** at *t*_4_) with an increasing relative attribution from persistent organ damage and dysfunction (**P1**).

#### 2.2.2. Scenario 2: Severe disease observed at t_1_

The model depicts a 5% probability of persistent organ dysfunction (**P1**), arising from observed severe COVID-19 (**V1**) and latent reversible organ dysfunction (**R1**, probability 100%).^4^ In this context the probability of reporting symptoms (**S1**) is 94%. The severity of the disease influences the probability of treatment (**T1**, probability 80%) which may precede functional recovery from *t*_2_ (**R2**, probability 13%) to *t*_4_ (**R4**, probability 3%). Similar to scenario 1, the probability of sustained reversible organ dysfunction drops to 13% at *t*_2_, 6% at *t*_3_ and 3% at *t*_4_, as the underlying tissue damage and dysfunction recovers. Meanwhile, the probability of persistent organ dysfunction (**P1**) stays relatively constant throughout the modelled follow-up period. The probability of reported symptoms also drops from 94% at *t*_1_ (**S1**) to 7% (**S4**) at *t*_4_.

#### 2.2.3. Scenario 3: Symptoms reported at t_1_, indicating acute COVID

If an adult reports symptoms (**S1**) at *t*_1_, and we suspect COVID-19 but have no other firm information about their disease status, the probability of having mild COVID-19 is 78% and the chance of progression to severe disease (**V1**) is 12%. In this context, the probability of symptoms arising as a result of underlying reversible organ dysfunction is high (**R1**, 98%) and decreases throughout the modelled follow-up period, alongside the declining probability of reporting symptoms at *t*_2_ (17%), *t*_3_ (9%), and *t*_4_ (7%).

#### 2.2.4. Scenario 4: Symptoms reported at t_1_ and t_3_, indicating long COVID

An adult reports symptoms at the time of acute infection (**S1**) but also beyond this time point, at *t*_3_ (**S3**). This scenario reflects many operational definitions of long COVID (including the definition used by Darley and colleagues in the ADAPT study), and is circumscribed to ongoing physical (e.g., dyspnoea, chest pain, fatigue/malaise) and/or neuropsychiatric symptoms lasting longer than 3 months after acute SARS-CoV-2 infection.[11] The probability of these symptoms being attributed to mild COVID-19 (**M1**) is 58%, lower than for scenario 3. However, the probability that this person had severe COVID-19 increases to 17% (compared to the 12% of scenario 3 [**V1**]).

Ongoing and recurrent symptoms (**S3**) at *t*_3_ result from persistent organ dysfunction, which may give rise to newly emerging symptoms (**ES3**) at the same time point (10% probability), and further ongoing and recurrent symptoms beyond *t*_3_ (**S4**, 52% probability).

### 2.3. Applying the general framework to the pulmonary system

The general framework can be used to organise and align proposed mechanisms for COVID-19-related dysfunction of the pulmonary system. The proposed mechanisms are generally latent (i.e., cannot be directly observed) and have included organ damage and dysfunction, persistence of the SARS-CoV-2, protracted immune dysregulation, microangiopathy, disseminated microthrombosis, and mitochondrial dysfunction. Here we select a small number of these hypothesised pathobiological mechanisms and illustrate how we can map them to the general framework to underpin causal inference and statistical analyses. Figure 4 presents the selected hypotheses aligned with the framework. The general node classes (such as **R1** and *S1*) have been replaced throughout with the appropriate specific latent and observable mechanisms (such as **alveolar inflammation** for **R1** and **cough** and **difficulty breathing** for **S1**). Not every possible mediating mechanism has been, nor need be, included in the mapping, as we explain below.

**Figure 4:**
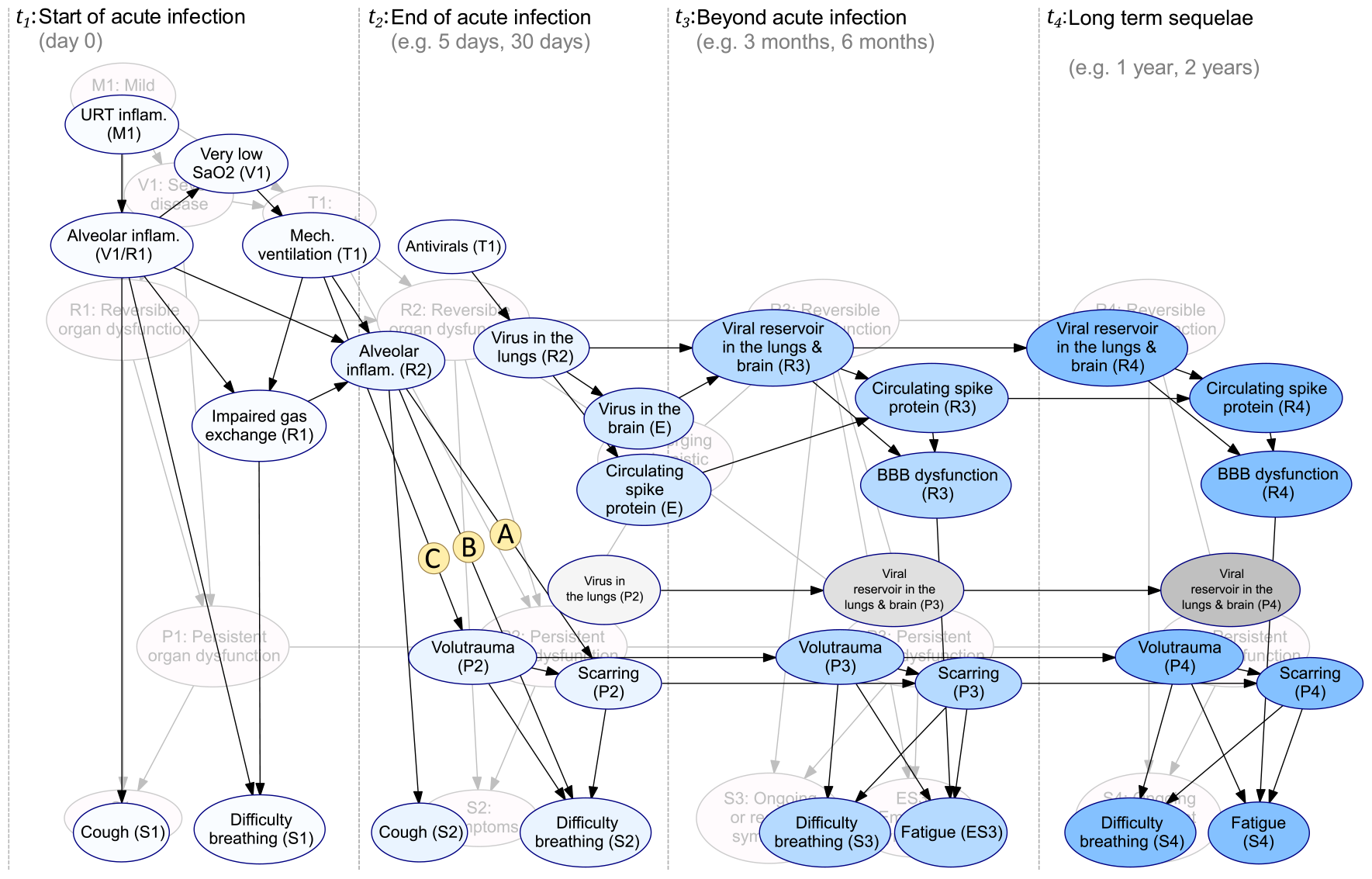
Application of the general framework to the pulmonary system, including initial SARS-CoV-2 infection, pulmonary symptoms, and the emergence of fatigue. The general framework is visible in the background, and the time points in this example remain aligned with those in Figure 2.

**Alveolar inflammation** is a prominent latent mechanism at *t*_1_ that can give rise to **cough** and **difficulty breathing**.[13] Of the many potential treatment options during *t*_1_, we have included **antivirals** (e.g., nirmatrelvir/ritonavir) and **mechanical ventilation** as examples. **Antivirals** may reduce pulmonary viral replication, potentially ameliorating more serious consequences caused by SARS-CoV-2 induced inflammation and tissue damage.[14] **Mechanical ventilation** may ensure the patient has an adequate pulmonary function, but can also lead to **volutrauma** (a type of ventilator-induced lung injury due to lung overdistension) with further release of pro-inflammatory cytokines that may induce or aggravate ongoing lung injury (arrow C), especially among those with pre-existing comorbidities like chronic obstructive pulmonary disease.[15]

When homeostatic mechanisms fail, transudative fluid can accumulate in the lower airways (alveoli) at *t*_2_ which can lead to the development of acute respiratory distress syndrome (ARDS). ARDS contributes to worsening of **difficulty breathing** and can result in chronic pulmonary inflammation and lung remodelling. In some instances, lung remodelling may progress to fibrosis (i.e., **scarring**, arrow A) owing to diffuse alveolar damage and the formation of alveolar hyaline membranes.[17] Additional pathological mechanisms involved in the development of ARDS and subsequent **scarring** include disseminated pulmonary micro-immunothromboses (*t*_2_, and possibly *t*_3_) leading to alveolar-capillary dysfunction, impaired gas exchange and hence, **difficulty breathing** and **fatigue**.[18] The extent to which these factors contribute to long-term respiratory outcomes may be compounded by additional factors, such as **volutrauma** at *t*_2_, *t*_3_, and *t*_4_ and emerging mechanisms.

It has been hypothesised that emerging mechanistic pathways (such as via **circulating spike protein**) contributing to long COVID could arise from the persistence of viral antigens or SARS-CoV-2 RNA in the respiratory epithelium (**virus in the lungs**) leading to prolonged immune activation, chronic pulmonary inflammation and persistent tissue damage and dysfunction [19]. Analogous mechanisms (such as immune activation and tissue damage) could emerge from the persistence of SARS-CoV-2 in the pulmonary and extrapulmonary tissues (e.g., **virus in the brain**).[2] Reservoirs of SARS-CoV-2 (**Viral reservoir in the lungs & brain**) that can increase circulating spike protein and further enhance endothelial and blood-brain barrier dysfunction (**BBB dysfunction**), leading to the emergence or worsening of preexisting COVID-driven **fatigue** at *t*_3_ and/or *t*_4_.[20] **Scarring** and **volutrauma** may compound the issue.

In the causal model, we have represented proposed neuropathological mechanisms for **fatigue** as reversible, running alongside **R2, R3**, and **R4**. However, it is possible that some mechanisms are persistent, and we have depicted this alternative hypothesis with a dashed outline. The general framework allows for both hypothetical forms to be represented and jointly tested, by (for example) showing emergence, maintenance and/or resolution of associated symptoms over the expected timeframes.

## 3. Discussion

Our aim was to help shape the big picture understanding of long COVID alongside research tools to facilitate efficient communication among research groups and those with lived experience of long COVID. We have applied a causal modelling approach to represent the proposed pathobiological mechanisms underpinning long COVID while accounting for uncertainties. We have demonstrated how this general causal BN framework can be used as a model to help explain how variation in study design might give rise to a range of estimates of long COVID prevalence and impact, allowing such studies to be compared. We have also shown how the general framework can be applied to depict current understanding of the complex pathology of long COVID involving the pulmonary system. In so doing, we believe we have demonstrated that this simple BN framework has practical value for research into long COVID.

Since a causal BN underpins the framework, it can be used to help address both diagnostic and prognostic needs in long COVID. In diagnostic problems (Figure 2) inference flows opposite to the direction of the network arrows from (observed) effects to their causes. *Reported* symptoms or clusters of symptoms (**S1** or **S2**) during the acute phase of COVID-19 (i.e., between *t*_1_ and *t*_2_) propagate backwards to influence the estimated probability of reversible or persistent organ damage and dysfunction (**R1, R2, P1**, and **P2**). Studies focusing on identifying the presence of long COVID or identifying potential endotypes of long COVID would find the diagnostic use of this framework valuable. For prognostication, by contrast, information propagates forward along causal links, following the direction of the network arrows. For example, mild COVID-19 (**M1**) has a direct causal effect on the probability of reversible organ dysfunction (**R1** and **R2** via **T1**) and the probability of receiving antiviral treatment (**T1**). This affects the probability of the subsequent development of symptoms (**S1**) which can be ongoing across the depicted follow-up period (**S2, S3, S4**), conditional on the patency of reversible (**R2, R3**, and **R4**) or persistent organ dysfunction (**P2, P3**, and **P4**). In this framework, these predictions also require the explicit account of interventions like antiviral treatments, allowing one to estimate the effects of one treatment compared to another, or to no treatment. As an explicitly causal framework, Pearl’s do-calculus can be applied to assess the effects of treatment, so long as the requirements of the calculus are adhered to, i.e., the known relevant background factors and confounders are also included.[21,22] The framework itself can help prognostic studies by clarifying what needs to be accounted for in the dynamics of long COVID. Prognostic applications, such as prediction tools and models, could also benefit from the ability to incorporate the diversity of existing studies in a framework that allows them to be unified.

In general, we see three major applications of the proposed causal BN framework. First, articulate a causal understanding of long COVID by applying the general framework to each specific aspect of interest (e.g., the pulmonary system, Figure 4), and by incorporating additional factors relevant to the research question of interest (such as COVID-19 vaccination, new types of SARS-CoV-2 strains, and newly identified disease mechanisms). Second, based on a clearer understanding, research studies can be designed to answer more explicit causal questions, which would inform the analysis of observational studies and trials, and how and when data should be captured (with project-specific time windows). Third, quantitative parametrisation of these networks, based on the results of existing quantitative studies that cover different parts of the network, could yield meaningful new insights into long COVID. Ultimately, such a parametrised network could inspire the discovery of latent relationships between symptoms and hypothesised disease mechanisms, and improve the prospects for predicting outcomes among those affected by long COVID.

One of the great difficulties that research into long COVID has faced is the uncertainty around what it and the set of associated conditions represent and their underlying mechanisms. These shortcomings have in turn led to difficulties in developing and agreeing to a common language. By making use of a knowledge engineering process in which we examined and validated the framework with domain experts, we were able to more clearly understand these problems and explicitly account for them. Our approach avoids specific definitions of LC and related conditions, but allows them to be mapped to a common framework based on a simpler and minimalist causal language. This also enabled us to perform initial validation of the generalisability of our approach to different clinical manifestations. As a consequence, we feel confident that the framework can provide a common language for the variety of syndromes that fall within the long COVID spectrum.

Finally, the proposed BN framework could be applied to better understand debilitating long-term phenomena such as post-infection myalgic encephalomyelitis (ME/CFS) as well as post-acute infection-specific syndromes (e.g., post-EBV or post-Ebola infections). Furthermore, the proposed causal modelling framework is expected to assist the design and analysis of observational studies as well as the selection of endpoints for randomised controlled trials. Likewise, the framework aims to facilitate the discovery and validation of treatment and diagnostic strategies for long COVID and associated phenomena.

## 4. Methods and Materials

We applied a knowledge engineering approach to create the causal models for long COVID (DAGs and BNs). The steps for developing each model followed a sequence summarised elsewhere.[13] The preliminary models and overall framework were drafted by the modelers based on published literature. These outputs were presented to clinical domain experts in infectious diseases (TS and GM, see Acknowledgements) and long COVID (GM) via a knowledge elicitation session to consolidate understanding of the problem domain. The framework was then refined and presented to an immunobiologist with expertise on long COVID (CP) via a validation session. The elicitation sessions were led by an experienced facilitator (SM).

As part of the validation session, we tested the applicability of the time frames chosen for the framework, explicitly considering their relevance to the ADAPT study,[11] the largest observational longitudinal study of Australian adults with PCR-confirmed SARS-CoV-2 and mild-to-moderate COVID-19.

To illustrate the ability of our causal modelling approach to handle common long COVID scenarios (discussed in the Results section) qualitative parameterisation was undertaken by the modellers for exploratory purposes using an intermediate causal structure.[10]. This technique uses plausible parameterisations to capture the qualitative behaviour of the system, such as the direction of changes in response to entering new information, without focusing on numerical precision.

## Data Availability

All data including source models and dictionaries are available on the Open Science Framework

https://osf.io/6q4h2/?view_only=3daeb277487c4ec39bee122dfd585baf

## 5. Acknowledgements

The authors are grateful for the input provided by Professor Gail Matthews (Program Head, Therapeutic Research and Vaccine Program at The Kirby Institute, University of New South Wales, Australia), Barbara Daniels and Belinda Saunders (consumer advisors from the Vaccines and Infectious Diseases Advisory Group at the Wesfarmers Centre of Vaccines and Infectious Diseases, Telethon Kids Institute, Nedlands, Western Australia, Australia).

## 7. Ethics

Ethics approval was granted by the Monash University Human Research Ethics Committee (Project ID 26942).

## 8. Appendix

Data dictionaries are available on the Open Science Framework.

## Footnotes

4 While latent variables would not normally have 100% posteriors, our definition of severe COVID-19 entails the presence of at least reversible organ dysfunction.

